# Toward Using Twitter for PrEP-Related Interventions: An Automated Natural Language Processing Pipeline for Identifying Gay or Bisexual Men in the United States

**DOI:** 10.1101/2021.08.23.21261924

**Authors:** Ari Z. Klein, Steven Meanley, Karen O’Connor, José A. Bauermeister, Graciela Gonzalez-Hernandez

**Author notes:** Corresponding author: Ari Z. Klein, 421A Blockley Hall, 423 Guardian Dr., Philadelphia, PA, USA 19104.

## Abstract

**Background:** Pre-exposure prophylaxis (PrEP) is highly effective at preventing the acquisition of Human Immunodeficiency Virus (HIV). There is a substantial gap, however, between the number of people in the United States who have indications for PrEP and the number of them who are prescribed PrEP. While Twitter content has been analyzed as a source of PrEP-related data (e.g., barriers), methods have not been developed to enable the use of Twitter as a platform for implementing PrEP-related interventions.

**Objective:** Men who have sex with men (MSM) are the population most affected by HIV in the United States. Therefore, the objective of this study was to develop and assess an automated natural language processing (NLP) pipeline for identifying men in the United States who have reported on Twitter that they are gay, bisexual, or MSM.

**Methods:** Between September 2020 and January 2021, we used the Twitter Streaming Application Programming Interface (API) to collect more than 3 million tweets containing keywords that men may include in posts reporting that they are gay, bisexual, or MSM. We deployed handwritten, high-precision regular expressions on the tweets and their user profile metadata designed to filter out noise and identify actual self-reports. We identified 10,043 unique users geolocated in the United States, and drew upon a validated NLP tool to automatically identify their ages.

**Results:** Based on manually distinguishing true and false positive self-reports in the tweets or profiles of 1000 of the 10,043 users identified by our automated pipeline, our pipeline has a precision of 0.85. Among the 8756 users for which a United States state-level geolocation was detected, 5096 (58.2%) of them are in the 10 states with the highest numbers of new HIV diagnoses. Among the 6240 users for which a county-level geolocation was detected, 4252 (68.1%) of them are in counties or states considered priority jurisdictions by the *Ending the HIV Epidemic (EHE)* initiative. Furthermore, the majority of the users are in the same two age groups as the majority of MSM in the United States with new HIV diagnoses.

**Conclusions:** Our automated NLP pipeline can be used to identify MSM in the United States who may be at risk for acquiring HIV, laying the groundwork for using Twitter on a large scale to target PrEP-related interventions directly at this population.

## Introduction

Pre-exposure prophylaxis (PrEP) with antiretroviral drugs is highly effective at preventing the acquisition of Human Immunodeficiency Virus (HIV) in men who have sex with men (MSM) [1]. There is a substantial gap, however, between the number of people in the United States who have indications for PrEP, including 25% of MSM [2], and the number of them who are prescribed PrEP [3]; approximately one third of primary care physicians (PCPs) in the United States who are aware of PrEP have prescribed PrEP or referred a patient for PrEP [4]. While efforts should be made to increase PCPs’ adoption of PrEP recommendations into routine clinical practice, PCP-based interventions are limited because some MSM, especially younger men, face challenges disclosing their same-sex sexual behaviors to their PCPs [5]. Based on the findings of Reuter et al.’s recent study [6] that examined Twitter users’ attitudes toward being monitored for health-related research, some MSM may be more open to PrEP-related interventions on social media, such as targeted messages or advertisements.

Hannaford et al. [7] found that social media can help identify factors for implementing PrEP-related interventions that are not captured by traditional research methods, and they suggest that, in turn, social media may present novel opportunities to implement PrEP-related interventions. While Twitter content has been analyzed as a source of PrEP-related data (e.g., barriers) [8,9], as far as we know, methods have not been developed to enable the use of Twitter as a platform for PrEP-related interventions. Implementing PrEP-related interventions on Twitter requires, foremost, identifying users in populations with indications for PrEP. Given that MSM are the population most affected by HIV in the United States [10], the objective of this study was to develop and assess an automated natural language processing (NLP) pipeline for identifying men in the United States who have reported on Twitter that they are gay, bisexual, or MSM. This study seeks to lay the groundwork for using Twitter on a large scale to target PrEP-related interventions directly at MSM who may be at risk for acquiring HIV.

## Methods

### Data Collection

The Institutional Review Board (IRB) of the University of Pennsylvania reviewed this study and deemed it exempt human subjects research under Category (4) of Paragraph (b) of the US Code of Federal Regulations Title 45 Section 46.101 for publicly available data sources (45 CFR §46.101(b)(4)).

Between September 2020 and January 2021, we used the Twitter Streaming Application Programming Interface (API) to collect more than 3 million tweets containing keywords that men may include in posts reporting that they are gay, bisexual, or MSM. As a preliminary approach, we deployed handwritten, high-precision regular expressions—search patterns designed to automatically match text strings—on the 3 million tweets to filter out noise and identify actual self-reports. After automatically removing retweets and “reported speech” (e.g., quotations, news headlines) [11], the regular expressions matched 8603 tweets, posted by 6358 users geolocated in the United States [12].

In addition to tweet-based regular expressions, we deployed handwritten regular expressions on the user profile metadata of the 3 million tweets collected from the Twitter Streaming API. The regular expressions matched the profile metadata of 4127 users geolocated in the United States [12]. After removing duplicate users from our tweet-based and profile-based searches, we identified a total of 10,043 unique users. Figure 1 illustrates our automated pipeline for identifying men in the United States who have reported on Twitter that they are gay, bisexual, or MSM. We analyzed the state- and county-level geolocations [12] of these 10,043 users, and drew upon a validated NLP tool [13] to automatically identify their ages.

**Figure 1.**
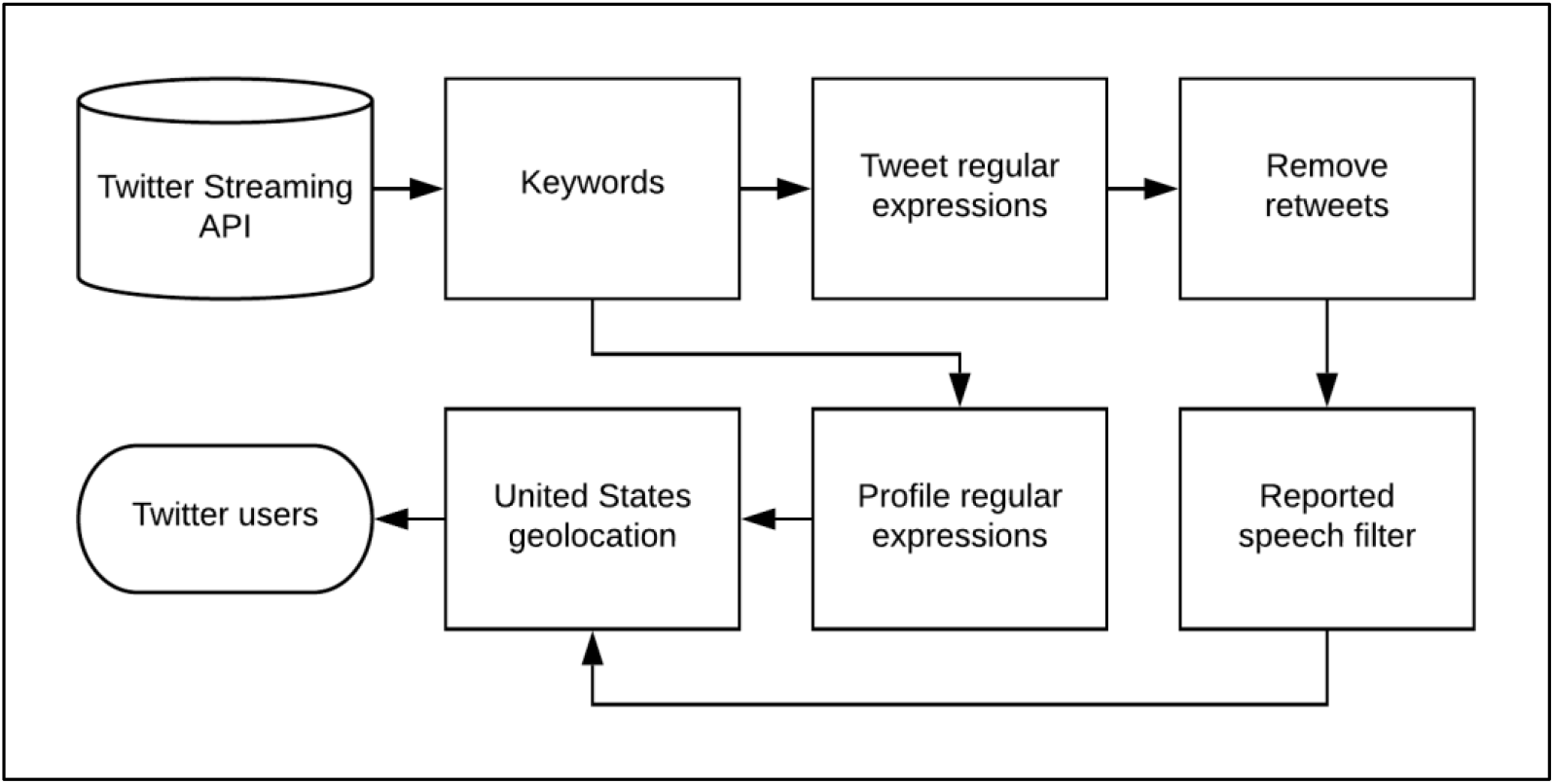
Automated natural language processing (NLP) pipeline for identifying men in the United States who have reported on Twitter that they are gay, bisexual, or men who have sex with men (MSM).

## Results

### Pipeline Validation

Two annotators manually distinguished true positives and false positives in a random sample of 1000 (10%) of the 10,043 users that were identified by our automated pipeline—500 of their matching tweets and 500 of their matching profiles—where *true positives* are tweets or profiles in which users report that they are gay, bisexual, or other MSM. Overall inter-annotator agreement (Cohen’s kappa), based on independent, dual annotations for all 1000 users, was 0.81, considered “almost perfect agreement” [14]. More specifically, inter-annotator agreement was 0.83 for the 500 tweets, and 0.79 for the 500 profiles. Upon resolving the disagreements, 417 (83.4%) of the tweets and 430 (86%) of the profiles were annotated as true positives, and 83 (16.6%) of the tweets and 70 (14%) of the profiles as false positives. Based on this evaluation, our automated pipeline has an overall precision of 0.85, where *precision = true positives / (true positives + false positives)*. Table 2 provides examples of tweets (T) and profiles (P) that were manually annotated as true positives (+) or false positives (-). The majority of profiles that were annotated as false positive report that the user is transgender or non-binary—populations beyond the scope of this particular study.

**Table 2.** Sample tweets (T) and profiles (P) that were identified by our automated natural language processing (NLP) pipeline and manually annotated as true positives (+) or false positives (-), where *true positives* are tweets or profiles in which users report that they are gay, bisexual, or other men who have sex with men (MSM).

| Type | Text | Label |
| --- | --- | --- |
| T | End the FDA’s discriminatory and unscientific policy against gay men like me donating blood. | + |
| T | As a bi guy we get so little representation, and almost all of its negative. It’s frustrating. | + |
| T | Today, we remember Matthew Shepard who’s life was cut short as a result of a hate crime due to his identity as a gay male. | - |
| P | A proud black gay guy. | + |
| P | 50+ gay trans man, writer, film and food lover. He/him OR they/them. | - |

### Demographics

We detected a United States state-level geolocation for 8756 (87.6%) of the 10,043 users identified by our automated pipeline, including users from all 50 states and the District of Columbia. As Figure 2 illustrates, the largest numbers of users were detected in California, New York, Texas, Florida, Illinois, Pennsylvania, Ohio, and Georgia. We detected a county-level geolocation for 6240 (71.2%) of these 8756 users. Figure 3 presents the 15 counties for which we detected at least 100 users. We detected an age of at least 13 years [10] for 4782 (47.6%) of the 10,043 users, with a mean age of 31.9, a standard deviation of 13.1, and a median of 29. Figure 4 presents the age distributions, based on each user’s most recent tweet containing a self-report of age.

**Figure 2.**
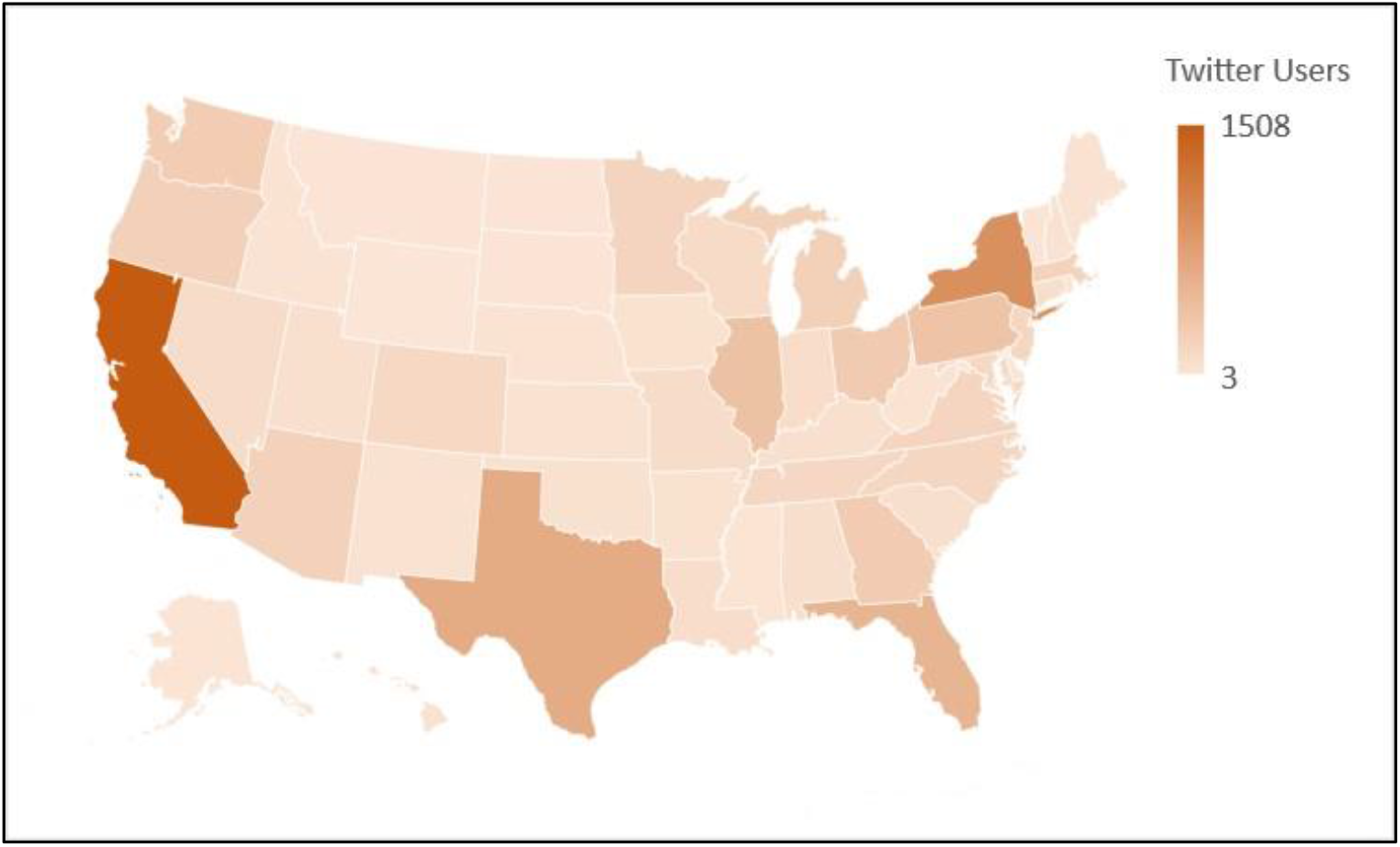
Men in the United States who have reported on Twitter that they are gay, bisexual, or other men who have sex with men (MSM), by state, between September 2020 and January 2021.

**Figure 3.**
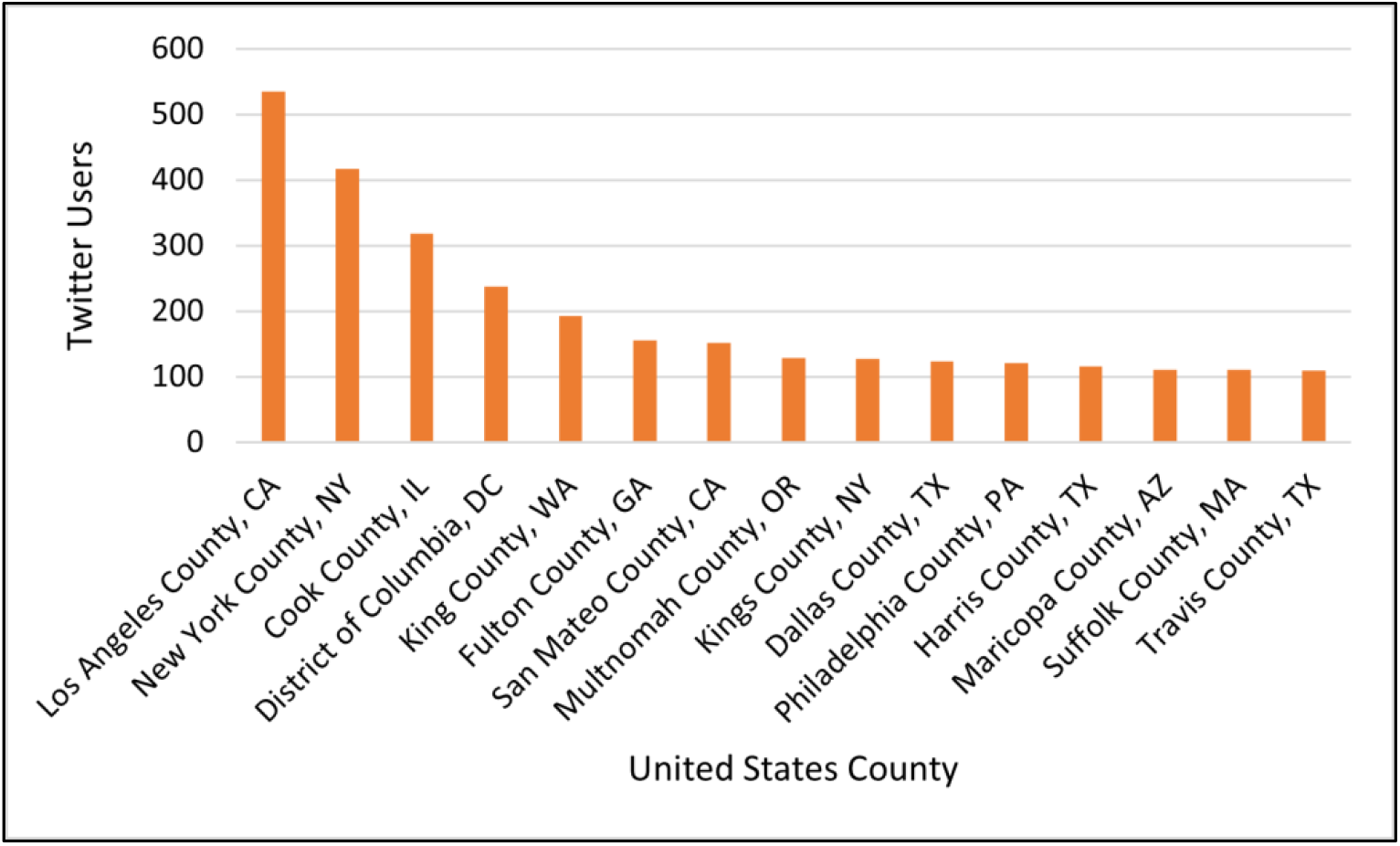
Men in the United States who have reported on Twitter that they are gay, bisexual, or other men who have sex with men (MSM), by county with at least 100 users detected by our automated pipeline, between September 2020 and January 2021.

**Figure 4.**
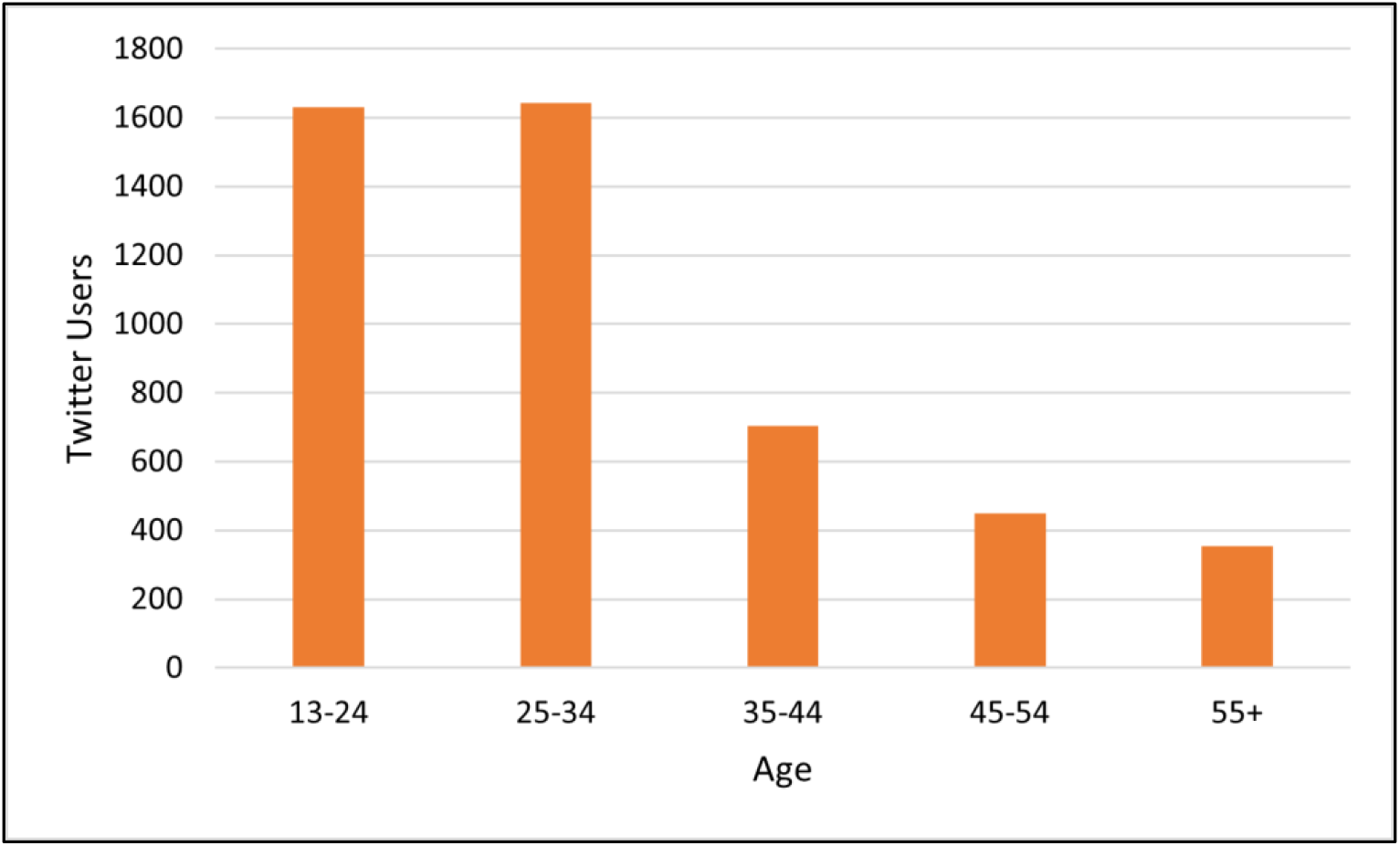
Men in the United States who have reported on Twitter that they are gay, bisexual, or other men who have sex with men (MSM), by age, between September 2020 and January 2021.

## Discussion

Our study demonstrates that gay, bisexual, or other MSM in the United States publicly report their sexual orientation on Twitter, and that these users can be accurately identified on a large scale. Moreover, among the 8756 users for which our automated pipeline detected a United States state-level geolocation, 5096 (58.2%) of them are in the 10 states with the highest numbers of new HIV diagnoses [10]. Among the 6240 users for which a county-level geolocation was detected, 4252 (68.1%) of them are in counties or states considered priority jurisdictions by the *Ending the HIV Epidemic (EHE)* initiative [15]. Furthermore, the distribution of the users’ ages reflects the ranking of age groups with new HIV diagnoses among MSM in the United States [10], with 25-34 first and 13-24 second; more specifically, the majority of the users are in one of these same two age groups as the majority of MSM with new HIV diagnoses [10]. The mean (31.9) and median (29) ages of the users are within the age range (25-34) with the largest number of new HIV diagnoses, which is also the only age range in which HIV infections have increased since 2014 [10]. Therefore, our automated pipeline can be used as the basis for PrEP-related interventions targeted directly at MSM who are largely in regions and age groups most affected by HIV in the United States, including younger men who may face challenges discussing their same-sex sexual behaviors with their PCPs [5].

## Conclusions

This paper presented an automated NLP pipeline that can be used to identify MSM in the United States who may be at risk for acquiring HIV, laying the groundwork for using Twitter on a large scale to target PrEP-related interventions directly at this population.

## Data Availability

The annotated Twitter data used to validate our automated natural language processing pipeline will be made available by request.

## Acknowledgments

AZK contributed to designing the pipeline, developing the sets of regular expressions, preparing the data set for validation, resolving the annotators’ disagreements, analyzing the demographics, and writing the manuscript. SM contributed to guiding the data collection from Twitter and the validation, and editing the manuscript. KO contributed to annotating the Twitter data for validation, calculating inter-annotator agreement, and editing the manuscript. JB contributed to guiding the overall study design and the data collection from Twitter, and editing the manuscript. GGH contributed to conceptualizing the research study, guiding the overall study design and the data collection from Twitter, and editing the manuscript. The authors would also like to thank Ivan Flores for contributing to software development, and Alexis Upshur for contributing to annotating the Twitter data for validation of the pipeline. This research was supported by a grant from the Penn Center for AIDS Research (CFAR), an NIH-funded program (P30 AI 045008).

